# Extension of Community Healthcare Outcomes in Parkinson Disease (Parkinson ECHO): A Feasibility Study

**DOI:** 10.1101/2022.01.31.22270212

**Authors:** Lee E. Neilson, Jennifer Wilhelm, Margaret McLain McDonnell, Lisa Mann, Jeff A. Kraakevik

## Abstract

Parkinson’s disease is the second most common neurodegenerative disorder and presents with a heterogeneous group of symptoms. Managing these symptoms requires coordinated care from a neurology specialist and a primary care provider. Access to neurology care is limited for those patients with Parkinson’s disease who reside in rural areas given financial and mobility constraints along with the rarity of specialty providers. To close this gap, we developed and implemented a telehealth-based Project ECHO^®^ (Extension for Community Healthcare Outcomes) program, “Parkinson ECHO,” to provide education and support for rural clinicians and allied health members. We assessed the feasibility of this tele-mentoring educational offering and the effect it had on clinician confidence in diagnosing and treating Parkinson’s disease. Participants from across Oregon (n=33), of whom 70% served rural and/or medically underserved communities, attended the biweekly sessions. The sessions focused on a topic within Parkinson’s disease diagnosis or management followed by case discussions. We assessed clinician confidence and acceptability using surveys before and after their participation in the program. The COVID-19 pandemic was an unexpected obstacle in executing this project, delaying the program by several months and likely accounting for a lower number of attendees for the last two sessions. Nevertheless, we found Parkinson ECHO did significantly increase participant confidence levels in diagnosing and managing Parkinson’s disease.

## Background

Parkinson’s disease (PD) is a neurodegenerative disorder characterized by rigidity, slowness of movement, tremor, and gait changes (Obeso *et al*., 2017). The pharmacological management of these motor symptoms has become increasingly complex (Armstrong and Okun, 2020). Less well-characterized are the non-motor symptoms, including depression, psychosis, constipation, sialorrhea and more. These non-motor symptoms are strongly associated with patient’s quality of life (Barone *et al*., 2009). Because of its heterogeneous presentation, PD requires multidisciplinary care. It has been shown that those patients with Parkinson’s disease (PwP) who have access to specialist care have lower risk of hospitalization for PD-related illnesses (Willis *et al*., 2012) and greater survival (Willis *et al*., 2011).

Yet access to specialist care is limited. While the supply of neurologists has remained relatively stable, the prevalence of neurologic disorders has substantially increased, leading to greater demand for neurology services. Moreover, there are great disparities between rural, medically-underserved regions and urban areas in terms of neurologist density (Lin *et al*., 2021). Thus, it is perhaps unsurprising that up to 40% of PwP in the United States do not see a neurologist (Willis *et al*., 2011). This may be one reason rural living is a significant negative predictor of health-related quality of life in PD (Soh *et al*., 2012).

In order to close this access gap, the Parkinson Foundation (PF), Oregon Extension for Community Healthcare Outcomes (ECHO) Network (OEN), and Oregon Health and Science University (OHSU) collaborated on a feasibility study to train rural and frontier providers based on the Project ECHO^®^ model. ECHO, is a hub-and-spoke system whereby a central academic ‘hub’ connects with the many spokes of primary care providers and allied health professionals throughout a large geographic area. By utilizing a model where participants educate each other and actively learn, these practitioners are better prepared to deliver higher quality care to patients with complex conditions.

The ECHO model rests on four pillars: 1) using technology (i.e. videoconferencing); 2) sharing of best practices; 3) case-based learning; and 4) monitoring of outcomes with regular surveys. While initially starting with hepatitis C in New Mexico (Arora *et al*., 2007), ECHO program topics have rapidly expanded to other chronic conditions. More recently it has expanded into the realm of neurological disorders including dementia (Lindauer *et al*., 2020), epilepsy (McDonald *et al*., 2021), and multiple sclerosis (Johnson *et al*., 2017, Alschuler *et al*., 2019).

To our knowledge, there is no published literature or existing program applying this model to PD. However, it is well known in PD that when generalist and specialist care reinforce one another, the PD patient benefits (Plouvier *et al*., 2017). Therefore, this ECHO model holds promise for improving the quality of life of PwP with limited access to specialists. In developing and piloting this project our objectives were 1) to evaluate the feasibility and sustainability of our program, 2) to evaluate the favorability of such a program; and 3) to evaluate the impact on clinician confidence in managing PD.

## Methods

### Overview and Program Development

The Parkinson ECHO team was multidisciplinary by design with a facilitator (RN/BSN), core faculty experts (MD, DPT), a scribe (MD), and guest speakers (MD, DPT). We devised six biweekly 1-hour sessions consisting of a welcome, roll call to facilitate name-face recognition, review of key concepts from the prior session (for sessions two through six), and a brief didactic led by a content expert (**Table 1**), with the remainder of the time devoted to case consultation. The sessions were augmented with online educational resources on our program website including patient education materials. Participants were invited to submit a recent encounter with a person with parkinsonism for shared problem solving on diagnosis and treatment options, with expert faculty facilitating. The session concluded with a brief summary of key points from the scribe, who also sent a written summary of the case recommendations one week later to reinforce learning points.

**Table 1.**

| Curriculum Topics and Dates |  |  |
| --- | --- | --- |
| Date | Topic | Presenter Credentials |
| 1/22/20 | Differential diagnosis of PD and related conditions | MD (neurology) |
| 2/5/20 | Treating motor and non-motor symptoms of PD | MD (neurology) |
| 2/19/20 | Safety concerns and improving team care | DPT and RN, BSN |
| 3/4/20 | Cognitive effects of PD, including depression and anxiety | MD (psychiatry) |
| *COVID-19 PAUSE* |  |  |
| 6/3/20 | Hospitalization and PD | MD (neurology) |
| 6/17/20 | Advanced stage PD and palliative care | MD (neurology) |

The program was scheduled at noon on Wednesdays, i.e. over the lunch hour, to maximize participant availability. The videoconferencing platform allowed participants to see each other as well as the presentation slides and images. A chat box feature was monitored, with encouragement to verbally participate. Technical expertise and onsite support was offered if needed.

### Protection of Human Subjects

This was submitted to the Institutional Review Board at OHSU as Study #00020683. They determined that the proposed activity is not research involving human subjects and that IRB review and approval was not required.

### Recruitment

In the recruitment phase of this project, we leveraged our partnership with OEN to recruit PCPs who care for PwP and were otherwise unaffiliated with a PD center of excellence for inclusion in this program.. Since this was primarily an educational offering, we did not exclude any interested party from participating.

### Outcome Measures

Within two weeks of starting the program, all participants were sent a web-based questionnaire regarding their clinical role, years of practice, number of PwP seen per month, knowledge questions to determine baseline understanding of PD-related issues, and additional questions surveying level of comfort with diagnosing and treating PD. Immediately after each individual session, participants were emailed a questionnaire for feedback on organization, relevance, use of evidence-based content and overall rating using a 5 point Likert scale. We made one follow up inquiry one week later to improve the response rate. Finally, after the 6 sessions, participants were asked to complete a post-training survey which included the original knowledge questions presented in the pre-program survey as well as open-field comment boxes for feedback. This was repeated 6 months later along with repeating surveys about level of comfort with diagnosis and treatment of PD. All surveys were sent by email through Research Electronic Data Capture (REDCap), a secure web application and database that is run through Oregon Clinical and Translational Research Institute (OCTRI). All participants were given an identification number to ensure confidentiality.

### Analysis

We did not perform a power analysis or have an enrollment target given the nature of this program. Demographic data and survey completion rates were tabulated and reported. Where appropriate, we performed paired 2-tailed t-tests with a significance level of 0.05 after confirming normality with Kolmogorov-Smirnov test given the small sample sizes.

## Results

Thirty-three unique participants from 13 Oregon counties and one county in the state of Washington signed up to participate (**Figure 1**). Of these, 14 self-identified as prescribing healthcare providers (MD = 5, DO = 1, NP = 6, PA = 2), with the remainder identified as naturopaths (1), physical therapists (2), occupational therapists (1), speech therapists (1), behavioral health specialists (3), registered nurses (3), with the remainder indicating ‘other’ or ‘no response’. Seventy percent (n=23) of participants reported serving rural and/or medically underserved populations. The prescribing providers reported seeing an average of 3 PwP on a monthly basis, with one reporting seeing greater than 13 PwP per month.

**Figure 1.**
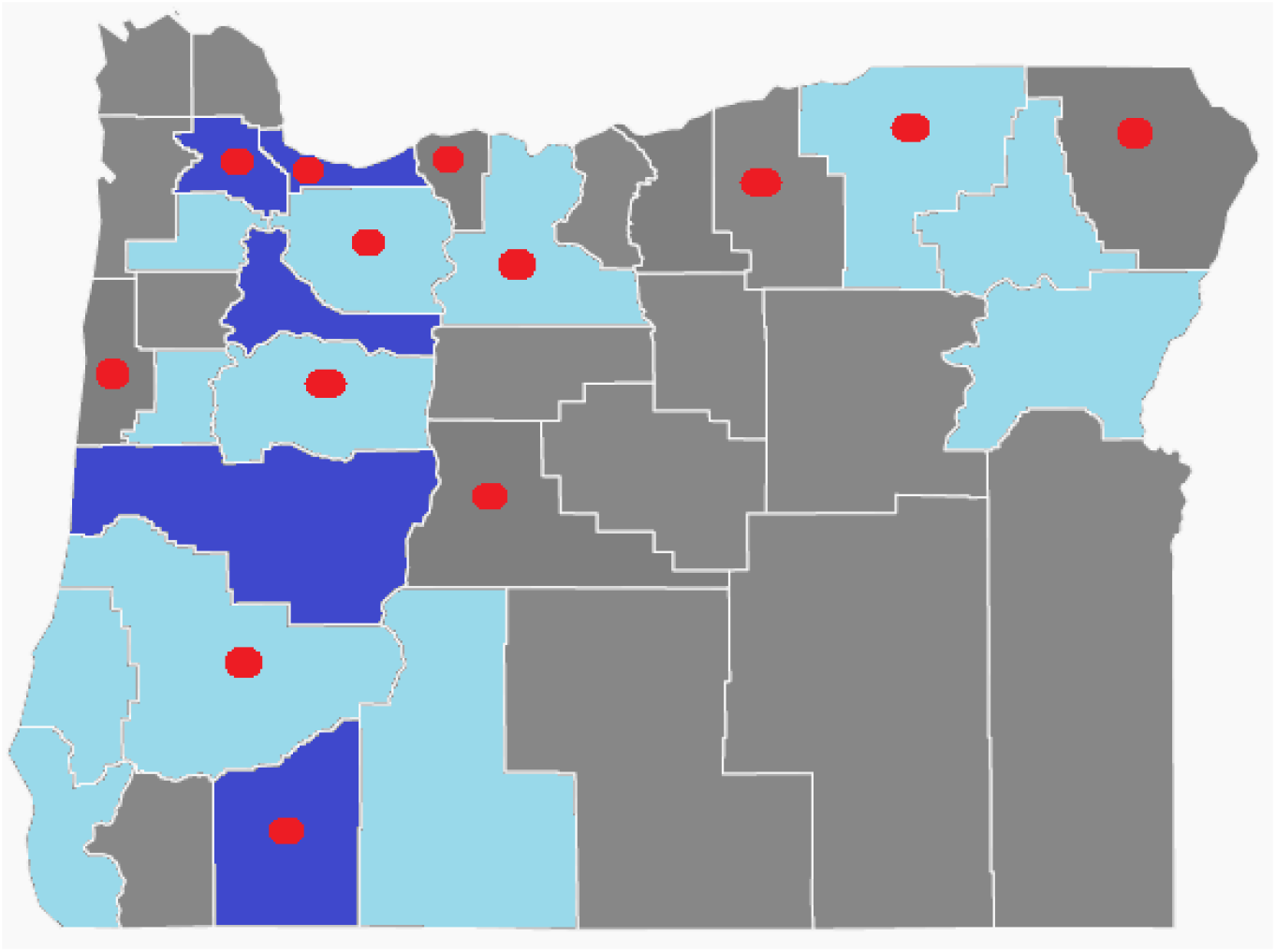
Map of the state of Oregon with county boundaries delineated. One participant came from Washington state and is not depicted. Dark blue = presence of movement disorder specialist; Light blue = general neurologist; Gray = absence of neurologist; Red = Parkinson ECHO participants

We planned for 6 biweekly sessions presented over 12 weeks; however, the COVID-19 pandemic interrupted our programming as healthcare systems shifted their focus to the evolving healthcare crisis. Thus, we completed the final two sessions after a three-month hiatus.

For the first four sessions, we averaged 22 participants (22, 28, 16, 21 in sessions 1 through 4, respectively), whereas 7 attended session 5 and 9 attended for the sixth and final session. In order to provide real-time feedback and to monitor for quality improvement, an online post-session survey was administered after each session using a simple 5-point Likert scale. As can be seen in **Table 2**, participants reported that the stated objectives were met (4.43/5) and that they were overall satisfied (4.44/5).

**Table 2.**

| Weekly Survey Responses |  |  |  |  |  |  |  |
| --- | --- | --- | --- | --- | --- | --- | --- |
| Question | Session 1<br>(n=18) | Session 2<br>(n=17) | Session 3<br>(n=13) | Session 4<br>(n=11) | Session 5<br>(n=6) | Session 6<br>(n=5) | Average Rating |
| Objectives met | 4.22 | 4.59 | 4.69 | 4.55 | 4.33 | 4.20 | 4.43 |
| Content was evidence-based | 4.11 | 4.53 | 4.69 | 4.55 | 4.33 | 4.20 | 4.40 |
| Pace | 4.17 | 4.41 | 4.69 | 4.45 | 4.50 | 4.20 | 4.40 |
| Organization | 4.33 | 4.65 | 4.77 | 4.73 | 4.67 | 4.20 | 4.56 |
| Relevance | 4.28 | 4.65 | 4.77 | 4.70 | 4.67 | 4.20 | 4.54 |
| Satisfaction | 4.17 | 4.65 | 4.77 | 4.45 | 4.60 | 4.00 | 4.44 |
| 1=poor, 2=fair, 3=good, 4=very good, 5=excellent |  |  |  |  |  |  |  |

In addition to weekly surveys, we also administered pre- and post-intervention surveys of Parkinson ECHO. Twenty six participants completed the pre-survey and 16 completed the post-survey, yielding response rates of 79% and 48% of total registered participants, respectively. Of the 16 participants who completed both the pre- and post-project surveys, we found significant improvement in comfort level in treating PD and a trend toward improved comfort in diagnosing PD. Of the 10 completers who indicated they had prescribing power, there was a significant improvement in comfort level in prescribing levodopa (**Table 2**).

On satisfaction measures, these participants all reported Parkinson ECHO was an effective format for learning and would recommend the program to a colleague. Supporting this quantitative data are select free-form quotes from participants indicating some examples of change in practice (**Table 4**).

**Table 3.**
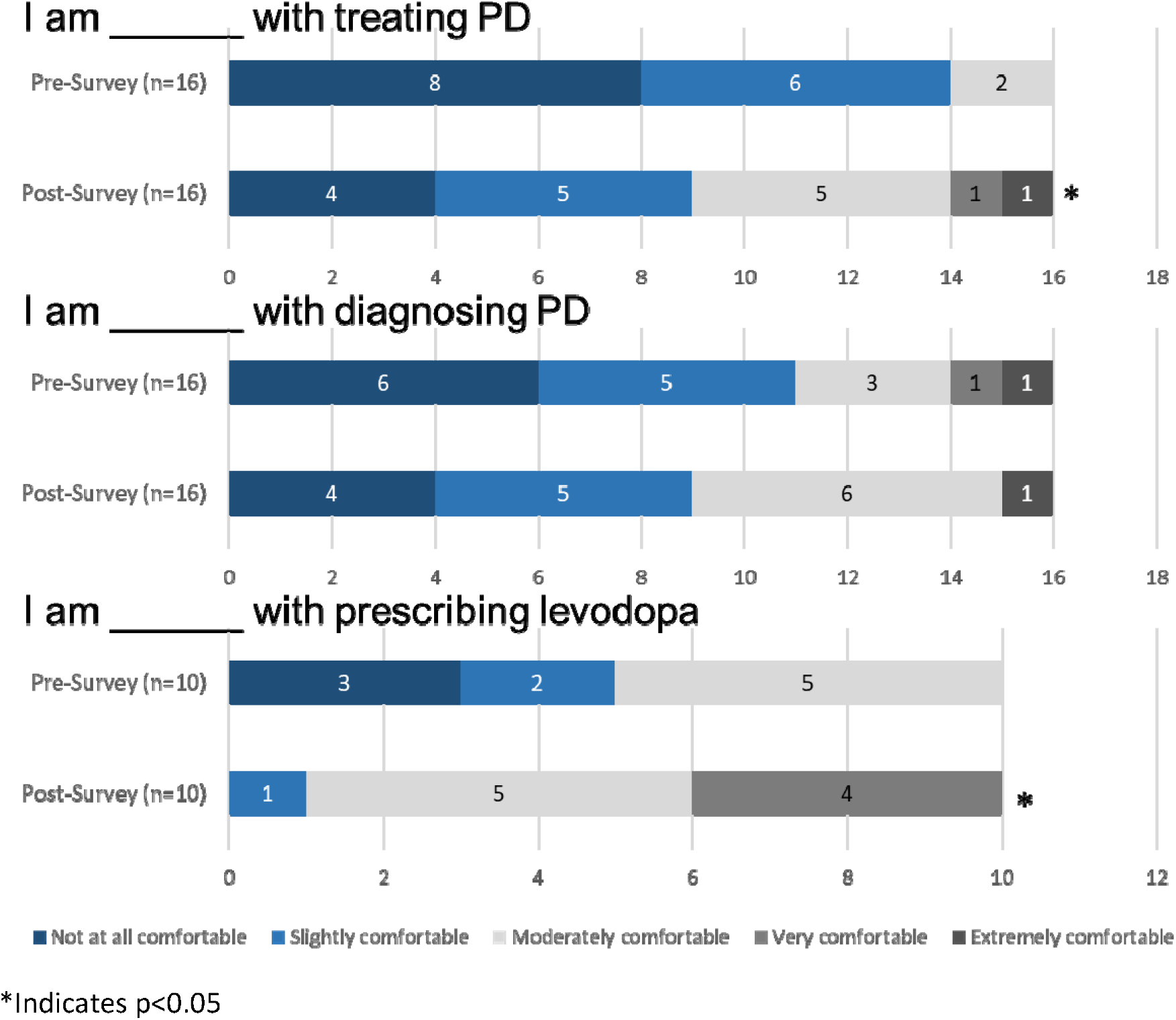

**Table 4.**
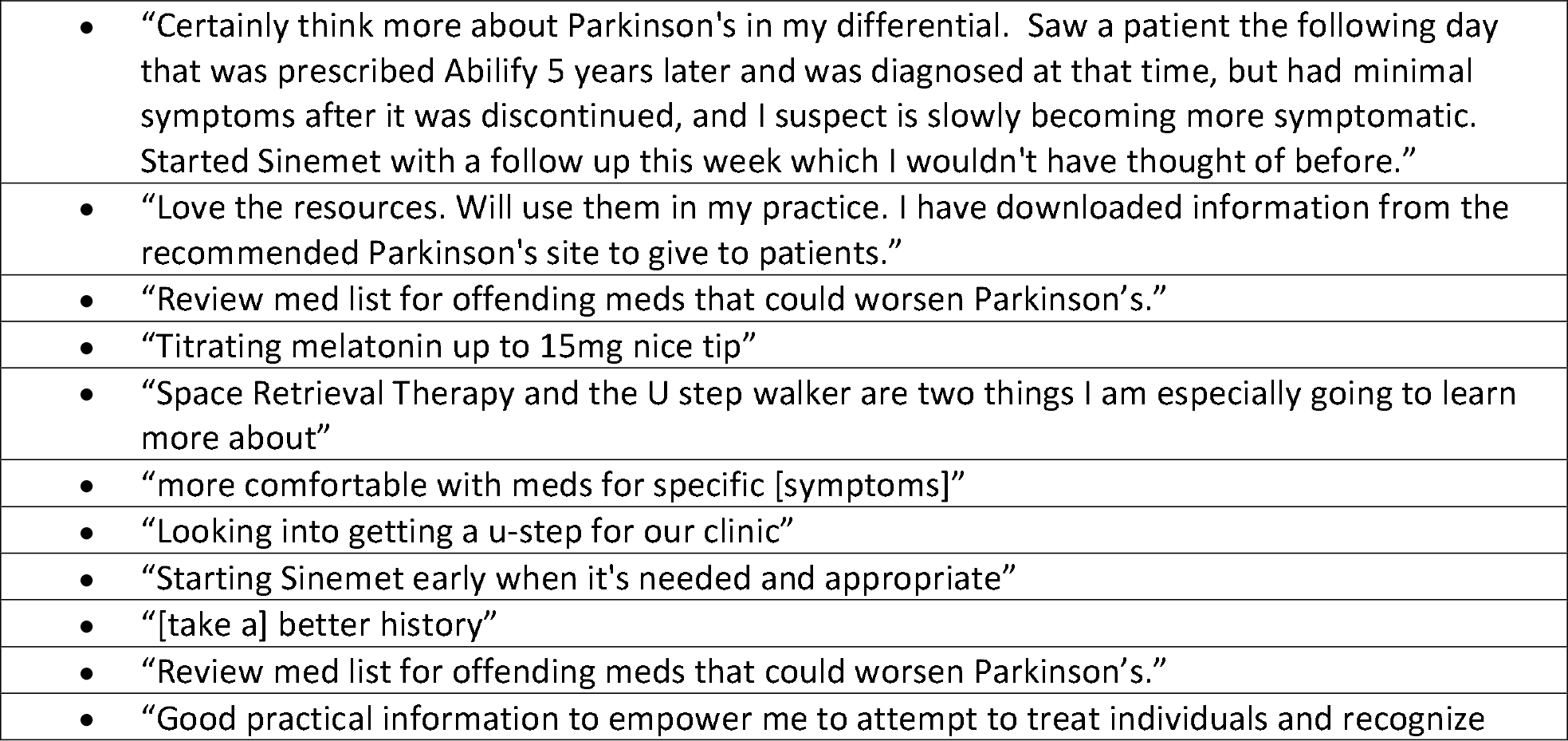
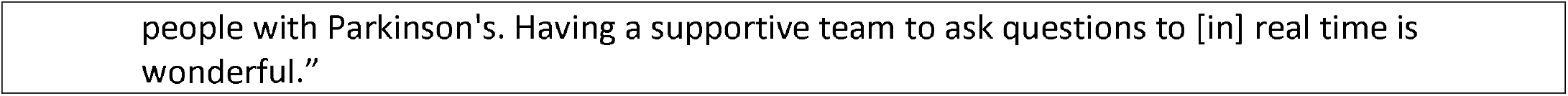
Selected feedback from participants, lifted directly, outlining practice changes made as a result of participation in ECHO-PD.

All 16 participants reported that Parkinson ECHO improved their satisfaction in treating PwPs, that learning from experiences of other primary care practices regarding PD was beneficial, and that participating in Parkinson ECHO is an effective way to enhance their clinic’s expertise. Furthermore, all 16 post-survey respondents reported that they would recommend Parkinson ECHO to a colleague after their experience in the program, and 7 of this sample expressed interest in face-to-face shadowing or ongoing mentorship at the academic hub. Encouragingly, two participants each reported they had convened a multidisciplinary group within their practice to discuss improving care and changed a policy, which led to change in patient care, and one participant led a didactic session for colleagues on PD.

## Discussion

To date, there have been limited studies targeting rural-dwelling PwP. A few models have been proposed including online health communities, nurse-led clinics, and telemedicine (Schuller *et al*., 2017). Online health communities do permit direct physician contact in addition to connecting with general patient resources, but patients may not feel empowered to get the specific help they need (Visser *et al*., 2016). One VA-based, nurse care manager-led program in the southwest United States could serve as a model to improve areas of need for PwP, but the training, resources, and infrastructure required to develop and maintain such a program may limit widespread use (Connor *et al*., 2019). Finally, telemedicine holds particular promise. One randomized controlled clinical trial compared those PwP with a stable internet connection to usual care with local physician or usual care plus four telehealth visits with a movement disorder specialist over 12 months. While they demonstrated a high completion rate and high satisfaction with the telemedicine format, there was no substantial change in quality of life measures, likely related to the high proportion of those already seeing a specialist as part of their usual care (Beck *et al*., 2017).

In contrast to these strategies, Parkinson ECHO, to our knowledge the first Project ECHO^®^ with a specific and primary focus on PD, attempted to reach rural-based primary care clinicians and allied health professionals. At the time of our program, there were 258 neurologists in the state of Oregon with an active medical license in neurology, of whom 213 (83%) practice in the six counties where a movement disorder specialist resided (Figure 1). An additional 12 counties have a licensed neurologist that does not specialize in movement disorders. These 12 counties, and the remaining 18 counties with no neurology presence, represent a large geographic area of underserved patients. We were able to reach 10 counties with no movement disorder presence, and overall, 70% of participants reported serving a rural population.

Despite the unplanned interruption due to COVID and the new challenges the faculty leaders and participants faced, the program was ultimately completed as planned. Therefore, not only is this format feasible, but planning is underway for an expanded second offering, suggesting it is sustainable. However, these offerings do often depend on grant funding or institutional support.

While it was intentional to reach primarily healthcare prescribers, given the multidisciplinary nature of PD care, it was encouraging that there was interest from several allied health professionals including those in behavioral health and rehabilitation services. Participants recorded high week-to-week satisfaction and all post-survey respondents reported recommending it to their colleagues. Perhaps most surprisingly, yet encouragingly, given the relatively small number of participants, 2 practices convened multidisciplinary groups to change practice.

In just a short time-frame, we were able to demonstrate significant changes in confidence and comfort level in diagnosing and treating PD. The only measure which did not achieve statistical significance, though still trended upward, was confidence in managing the hospitalized PD patient. There are a few explanations for this. First, the session explicitly dedicated to this topic came following the pause and had low attendance. Second, Parkinson ECHO was designed to improve outpatient care; those with inpatient responsibilities likely are fewer in number, and the inpatient setting may come with several checks in place. For example, it is common for electronic health records to alert to possibly dangerous drug-drug interactions. Future iterations could be designed to encompass more inpatient care.

Of the 6 cases presented by outlying participants, some common themes emerged. They included medication management (n=6), diagnosis (n=4), falls (n=3), caregiver burden (n=3), dementia (n=3), psychosis (n=4), sleep (n=2), and constipation (n=1). While each of the first four themes could be matched one-to-one with our planned didactics, the remaining non-motor symptoms proved to be a major point of focus by the participants. Given these non-motor symptoms are a strong driver of quality of life and are less often recognized as sequelae of PD outside of specialized PD centers, it would be prudent to expand this topic in future iterations.

Our pilot program does have some additional limitations. First, it is a pre– and post-study with no comparison group, so it is possible that knowledge and practice changes were due to other causes. However, our qualitative evaluation asked specifically about participants’ experiences, lessons, and practice changes resulting from Parkinson ECHO to mitigate this confounder. Second, we describe self-reported provider-level outcomes, which may be subject to bias. We emphasized the pre- and post-surveys where we had complete data, but obtaining weekly responses was difficult and could be an undue burden on a busy clinician. We did not measure patient-level outcomes. Future work should explore objective provider-level outcomes, perhaps through a chart audit evaluating the AAN practice parameters for PD (Chou *et al*., 2021), and patient experiences associated with provider participation in Parkinson ECHO.

Finally, our study lacks long-term follow-up. It is possible that the learning effect wanes overtime. It is also possible that a 6-session Parkinson ECHO, particularly one interrupted by a pandemic, is unable to detect changes in hospital policies or for providers to adapt their practice. However, the pandemic offered two additional strengths. First, in addition to reinforcing key concepts at the start of each session, the longer delay allowed a greater time lapse between sessions and inadvertently created a built-in spaced-repetition model of instruction. Second, the prolonged delay to collecting data allowed us to capture practice changes which typically require more time to enact following the conclusion of an educational program.

## Conclusion

Giving the disparity in outcomes, improving access to specialist care for PwP is a priority and we demonstrate that Parkinson ECHO can deliver the content effectively to rural and frontier clinicians who practice in counties without access to neurology care. Baseline preparedness and confidence in PD-related skills was low even among those motivated to attend. At the conclusion of the program, participants’ self-reported confidence in diagnosing and managing PD improved significantly. There were also several specific practice changes and two participants event convened multidisciplinary groups to enact larger scale change. This feasibility study should be followed up with replication and an assessment of objective quality metrics including review of patient records to evaluate for true practice change and connection of patients with resources.

## Data Availability

The raw data supporting the conclusions of this article will be made available by the authors, without undue reservation, to any qualified researcher.

## Author Contributions

LN prepared the figures and drafted the manuscript for intellectual content. LN and JW performed statistical analysis. All the authors contributed intellectually to the program and drafting of this manuscript. All authors read and approved the submitted version.

## Acknowledgments

The authors wish to thank Taylor Jay, PhD for providing constructive feedback on this manuscript.

## References

Alschuler, K.N., Stobbe, G.A., Hertz, D.P., Johnson, K.L., Von Geldern, G., Wundes, A., Reynolds, P., Unruh, K. & Scott, J.D., 2019. Impact of Multiple Sclerosis Project ECHO (Extension for Community Healthcare Outcomes) on Provider Confidence and Clinical Practice. International Journal of MS Care, 21, 143–150.

Armstrong, M.J. & Okun, M.S., 2020. Diagnosis and Treatment of Parkinson Disease: A Review. JAMA, 323, 548–560.

Arora, S., Thornton, K., Jenkusky, S.M., Parish, B. & Scaletti, J.V., 2007. Project ECHO: Linking University Specialists with Rural and Prison-Based Clinicians to Improve Care for People with Chronic Hepatitis C in New Mexico. Public Health Reports, 122, 74–77.

Barone, P., Antonini, A., Colosimo, C., Marconi, R., Morgante, L., Avarello, T.P., Bottacchi, E., Cannas, A., Ceravolo, G., Ceravolo, R., Cicarelli, G., Gaglio, R.M., Giglia, R.M., Iemolo, F., Manfredi, M., Meco, G., Nicoletti, A., Pederzoli, M., Petrone, A., Pisani, A., Pontieri, F.E., Quatrale, R., Ramat, S., Scala, R., Volpe, G., Zappulla, S., Bentivoglio, A.R., Stocchi, F., Trianni, G. & Dotto, P.D., 2009. The PRIAMO study: A multicenter assessment of nonmotor symptoms and their impact on quality of life in Parkinson’s disease. Movement Disorders, 24, 1641–1649.

Beck, C.A., Beran, D.B., Biglan, K.M., Boyd, C.M., Dorsey, E.R., Schmidt, P.N., Simone, R., Willis, A.W., Galifianakis, N.B., Katz, M., Tanner, C.M., Dodenhoff, K., Aldred, J., Carter, J., Fraser, A., Jimenez-Shahed, J., Hunter, C., Spindler, M., Reichwein, S., Mari, Z., Dunlop, B., Morgan, J.C., Mclane, D., Hickey, P., Gauger, L., Richard, I.H., Mejia, N.I., Bwala, G., Nance, M., Shih, L.C., Singer, C., Vargas-Parra, S., Zadikoff, C., Okon, N., Feigin, A., Ayan, J., Vaughan, C., Pahwa, R., Dhall, R., Hassan, A., Demello, S., Riggare, S.S., Wicks, P., Achey, M.A., Elson, M.J., Goldenthal, S., Keenan, H.T., Korn, R., Schwarz, H., Sharma, S., Stevenson, E.A. & Zhu, W., 2017. National randomized controlled trial of virtual house calls for Parkinson disease. Neurology, 89, 1152–1161.

Chou, K.L., Martello, J., Atem, J., Elrod, M., Foster, E.R., Freshwater, K., Gunzler, S.A., Kim, H., Mahajan, A., Sarva, H., Stebbins, G.T., Lee, E. & Yang, L., 2021. Quality Improvement in Neurology. Neurology, 97, 239–245.

Connor, K.I., Cheng, E.M., Barry, F., Siebens, H.C., Lee, M.L., Ganz, D.A., Mittman, B.S., Connor, M.K., Edwards, L.K., Mcgowan, M.G. & Vickrey, B.G., 2019. Randomized trial of care management to improve Parkinson disease care quality. Neurology, 92, e1831–e1842.

Johnson, K.L., Hertz, D., Stobbe, G., Alschuler, K., Kalb, R., Alexander, K.S., Kraft, G.H. & Scott, J.D., 2017. Project Extension for Community Healthcare Outcomes (ECHO) in Multiple Sclerosis. International Journal of MS Care, 19, 283–289.

Lin, C.C., Callaghan, B.C., Burke, J.F., Skolarus, L.E., Hill, C.E., Magliocco, B., Esper, G.J. & Kerber, K.A., 2021. Geographic Variation in Neurologist Density and Neurologic Care in the United States. Neurology, 96, e309–e321.

Lindauer, A., Wild, K., Natonson, A., Mattek, N., Wolf, M., Steeves-Reece, A. & Messecar, D., 2020. Dementia 360 ECHO: Using technology to facilitate diagnosis and treatment. Gerontol Geriatr Educ, 1–7.

Mcdonald, S.B., Privitera, M., Kakacek, J., Owens, S., Shafer, P. & Kobau, R., 2021. Developing epilepsy training capacity for primary care providers using the project ECHO telementoring model. Epilepsy Behav, 116, 107789.

Obeso, J.A., Stamelou, M., Goetz, C.G., Poewe, W., Lang, A.E., Weintraub, D., Burn, D., Halliday, G.M., Bezard, E., Przedborski, S., Lehericy, S., Brooks, D.J., Rothwell, J.C., Hallett, M., Delong, M.R., Marras, C., Tanner, C.M., Ross, G.W., Langston, J.W., Klein, C., Bonifati, V., Jankovic, J., Lozano, A.M., Deuschl, G., Bergman, H., Tolosa, E., Rodriguez-Violante, M., Fahn, S., Postuma, R.B., Berg, D., Marek, K., Standaert, D.G., Surmeier, D.J., Olanow, C.W., Kordower, J.H., Calabresi, P., Schapira, A.H.V. & Stoessl, A.J., 2017. Past, present, and future of Parkinson’s disease: A special essay on the 200th Anniversary of the Shaking Palsy. Movement Disorders, 32, 1264–1310.

Plouvier, A.O.A., Olde Hartman, T.C., Verhulst, C.E.M., Bloem, B.R., Van Weel, C. & Lagro-Janssen, A.L.M., 2017. Parkinson’s disease: patient and general practitioner perspectives on the role of primary care. Family Practice, 34, 227–233.

Schuller, K.A., Vaughan, B. & Wright, I., 2017. Models of Care Delivery for Patients With Parkinson Disease Living in Rural Areas. Fam Community Health, 40, 324–330.

Soh, S.E., Mcginley, J.L., Watts, J.J., Iansek, R. & Morris, M.E., 2012. Rural living and health-related quality of life in Australians with Parkinson’s disease. Rural Remote Health, 12, 2158.

Visser, L.M., Bleijenbergh, I.L., Benschop, Y.W.M., Van Riel, A.C.R. & Bloem, B.R., 2016. Do online communities change power processes in healthcare? Using case studies to examine the use of online health communities by patients with Parkinson’s disease: Table 1. BMJ Open, 6, e012110.

Willis, A.W., Schootman, M., Evanoff, B.A., Perlmutter, J.S. & Racette, B.A., 2011. Neurologist care in Parkinson disease: A utilization, outcomes, and survival study. Neurology, 77, 851–857.

Willis, A.W., Schootman, M., Tran, R., Kung, N., Evanoff, B.A., Perlmutter, J.S. & Racette, B.A., 2012. Neurologist-associated reduction in PD-related hospitalizations and health care expenditures. Neurology, 79, 1774–1780.

